# Health Worker Practices, Facilitators and Barriers towards the Intensified Case-Finding Tuberculosis Screening Tool in Uganda

**DOI:** 10.1101/2020.03.26.20043596

**Authors:** Michael Kakinda, Joseph K. B Matovu, Ekwaro A. Obuku

## Abstract

**Introduction:** Although the Tuberculosis (TB) Intensified Case Finding (ICF) tool was introduced in 2011, only 15.7% of the estimated 35 million people living with HIV were screened for TB in 2013. We explored the facilitators, barriers and health care workers’ practices regarding use of the ICF tool in TB screening.

**Methods:** We conducted a qualitative study in Jinja, eastern Uganda. We purposively sampled eight (4 private and 4 Government) health facilities (HFs) with the guidance of the District TB and Leprosy Supervisor (DTLS). At each health facility, three health care workers (in-charge TB clinic, a TB focal person & one laboratory technician (total: 24 participants in 8 HFs) were interviewed using a key informant interview guide. Data were collected on how TB was screened and diagnosed in general and when using the ICF tool in particular. Data were audio-recorded, transcribed in verbatim, coded and analyzed using a thematic framework.

**Results:** The ICF tool was available in all the 8 HFs; however, only half (12/24) the health workers interviewed at these facilities had ever used it for screening TB. The facilitators to ICF use were all levels of health cadres could use it, with simple, close-ended questions and clear, simple instructions. However, several barriers were identified as hindering the use of the ICF tool. The barriers to the use of the ICF tool are segmented according to the Health System building blocks, Leadership and Governance Barriers (concurrent use of other tools, lack of detailed training), Health Workforce Barriers (Lack of awareness of about the tool, perceived increased workload) and Health Information Management System Barriers (Stock-outs of the ICF tools).

**Discussion:** The ICF tool was found to be simple and easy to use; however, its use remained low due to a variety of perceived barriers by health workers. There is a need to increase the health care workers’ awareness about the ICF tool to improve its utilization in TB screening.

## Background

The World Health Organization (WHO) recommends regular screening for TB using the Intensified Case Finding Tool (ICF) especially among persons co-infected with HIV [1, 2]. Intensified TB case finding and prompt treatment aims to interrupt disease transmission and reduce TB associated morbidity and mortality [3]. ICF is the central intervention of the 3I’s strategy (Intensified Case Finding, Isoniazid Prevention Therapy and Infection Control), because it aims to identify patients as either having active TB (and in need of treatment) or being free of TB disease (and warranting preventive therapy) [4]. Also, Intensified Case Finding aides TB Infection control measures [5] as people who are likely to have TB can be separated from other patients.

While the use of the ICF tool improves TB screening [6], in 2013, only 5.5 million (15.7%) of the estimated 35 million people living with HIV [7] were screened for TB [8]. This suggests that the use of the ICF tool has not yet reached the optimal level and many TB suspected cases continue to interface with the health system in most developing countries but miss being detected and put on appropriate treatment. This is the first study of its kind, which tries to explain why the ICF tool is not being used as well as how it is being used. We explored the health workers’ practices regarding the use of the ICF tool in TB screening among health care workers in Uganda. We aimed to document the health system barriers and facilitators for ICF use in screening TB.

## Methods and Materials

### Study Setting

This study was carried out in Jinja Municipality, which is located in the eastern part of Uganda on the shores of Lake Victoria, 85 kilometres from Kampala, the Capital City of Uganda. Jinja District has a population of 468,256 people as of 2014 [9]. The district has 41 Government dispensaries (Health Centre IIs), 13 Health Centre III’s at the county level, 4 Health Centre IV’s at the sub-district level and one regional referral hospital. Also, there are 9 private or non-governmental organization dispensaries, 26 clinics, 3 Health Centers and 1 private hospital [10].

Among the government facilities, 3 offered general health services, one was a specialized hospital and a referral for the region with a special TB Clinic day every month. Two of the four 114 non-governmental health facilities were specialized clinics providing comprehensive HIV and TB care and diagnosis. The other 2 provided general health services.

#### Study Design, sampling and respondents

This was a qualitative case study using key informant interviews, participant observation and document analysis. We purposively selected eight health facilities (state and non-state) and 12 health workers (clinical, laboratory and management) in Jinja Municipality to achieve maximum variation. The health facilities were selected with guidance from the District TB and Leprosy Supervisor (DTLS). The respondents comprised three (3) health care, workers (in-charge TB clinic, a TB focal person & one-laboratory personnel) from each of the eight health facility for a total of 24 health care workers.

#### Data Collection Methods and Procedures

We collected data between March and July 2017. We conducted key informant interviews with health workers using a key informant interview guide. The guide was pre-tested among health workers of an HIV/AIDS clinic in Jinja municipality and adjustments were made. MK conducted the interviews in English in private consultation rooms. Participants described how they screened and diagnosed TB, including how they used the ICF tool. We probed for their perceptions of potential barriers or facilitators for using the ICF tool as well as health care workers’ practices regarding the use of the ICF tool for Tuberculosis screening. The interviews were audio-recorded with permission from the participants.

### Data Analysis

We transcribed the audio-recorded data from key informant interviews verbatim. We analyzed on a rolling basis for the validation of our findings. In subsequent analyses, we identified and coded data based on a priori themes including facilitators, barriers and health workers’ practices regarding the use of the ICF tool. MK and EAO read the transcripts to get an overview and subsequently identified units of analysis. MK and EAO agreed on the list of codes and then MK coded the material. All authors participated in condensing and summarizing the content of each code to make generalized descriptions concerning the use of the ICF tool.

### Ethics Approval

The Institutional Review Board (IRB) and the Faculty Review Board cleared the study for Health Sciences at Uganda Christian University. Informed consent was obtained from the participants in writing.

## Results

### Characteristics of respondents

The study participants were 24 health care workers. Their average age was 38 years (26-52), most of them were female (67%) while the rest were male. Most of the participants had diplomas in medicine (n=8), nursing (n=2) or medical laboratory technology (n=6). Only 2 of the participants had bachelor’s degrees (public health and medical laboratory technology) while the rest had certificates. All the participants had been practicing for a median of 5 years (IQR 3-13 years). ICF tools were available at all 8 participating health facilities. Of the 24 health workers interviewed, only 12 (50%) had ever used an ICF tool to conduct TB screening; 4 (17%) had ever seen it but never used it, while 8 (33%) had never seen it.

### Facilitators of the ICF Tool

Of those that had ever used the ICF tool, the main facilitator was the ease of its use because of clear instructions and simple language. Participants reported that the ICF tool had simple, closed-ended questions and was very brief thus requiring little time and effort to administer.

*“…The questions are easy to comprehend. You just tick. There are also a few fields to fill so it doesn’t take lots of time…”* Clinical Officer

*“*…*The questions of The ICF tool are straight forward so it can be used by anyone even a lay-person, it is also easy to use since it is a matter of just ticking*…*”* Enrolled Nurse

### Barriers to the use of the ICF Tool

The barriers to the use of the ICF tool are segmented according to the Health System building blocks, Leadership and Governance Barriers (concurrent use of other tools, lack of detailed training), Health Workforce Barriers (Lack of awareness of about the tool, perceived increased workload) and Health Information Management System Barriers (Stock-outs of the ICF tools).

### Leadership and Governance Barriers

Those that had never used the tool cited some challenges including the fact that there were other competing tools to complete, including tools used in other intervention such as prevention of mother to child transmission (PMTCT), early infant diagnosis of HIV (EID) and the integrated management of childhood illnesses (IMCI). Besides, recording individual patient data in the various registers was a challenge.

*“… There are many tools for all the various programs say PMTCT, IMCI, EID, and TB/Leprosy mention it so recording in a number of these tools is a challenge…”* Clinical Officer

Because the tool was assumed to be easy to use, the National TB and Leprosy Program and its partners decided that no detailed training was required and that all that was needed was an orientation during support supervision. This lack of knowledge now presents as a barrier to the use of the ICF tool. This finding suggests that even when the ICF tool orientation was provided, the message did not trickle down through to the health workers from the facility managers.

### Health Workforce Barriers

Lack of awareness about the tool was a major barrier; some health workers had never seen the ICF tool while others had only seen a sample during a TB/HIV collaborative workshop. It was due to these gaps that some health workers who did not understand the tool just ignored it. While the health workers at Out –Patient Department (OPD) and HIV clinics were knowledgeable about the tool and/or its purpose, most of the health workers in the Laboratory was often clueless.

*“…I don’t know it, but it is in the office of the in-charge, they used to send some people with the ICF tool but now they use their books for recording purposes…”*

Laboratory Technician.

*“… I think I do not know that tool I think you better brief me. The one for coughing for how long, night sweats, and the like, it is used on OPD but not in the Laboratory. Since we handle samples” …*Laboratory Technologist

Due to increased workload, the filling of the ICF tool was seen as an additional burden, as emphasized by the following quotation:

*“…The other barrier is some health workers are lazy to use them (ICF Tool) because of the workload”* … Enrolled Nurse

### Health Information Management System Barriers

The other barrier was that the ICF tool was said to be available in some facilities and not in others. It was reported to be available in facilities that did not use the tool. Even then, the facilities that reported using the tool also reported frequent stock-outs a few weeks after the supply of the ICF tool.

*“…Currently, we don’t have them but we used to have them. It was at every clinical room on the table. That was like 3 months ago and we were using them. It was here for a short time…”* Clinical Officer

Health care worker practices regarding the use of the ICF tool in the diagnosis and screening for TB

In four of the eight facilities, health workers reported using the ICF tool for symptomatic TB screening occasionally; irrespective of ownership status. Here, the ICF tool was often used at entry points especially in the Out-Patient Department (OPD), the HIV AIDS clinics and the Maternal Child Health Clinics. This was rarely the case in admission units. Hardly was the ICF tool employed in Health Centers levels III and IV.

Regarding how the tool was used, health workers reported that they mostly asked about a cough, which had lasted for a long period. Symptomatic patients were sent to the laboratory for sputum examination by microscopy. Priority was given to patients who had taken various antibiotics with no improvement.

*“… We use it when the patient has a cough for a long period. We administer it in the Out-Patient Department (OPD), we give it to people we suspect to have TB. Ideally, we should use it on patients who are on HAART (antiretroviral therapy) who have had a cough for over 2 weeks…’’* Clinical Officer

*“… TB is screened initially by using health education; the patient is asked about key signs and symptoms of Tuberculosis with the help of the ICF tool like cough for > 2 weeks if their sputum is blood-stained if they have noticed some weight loss. These clients will be isolated and taken to the TB waiting area. The other methods used for screening are requested by the clinician like the chest radiographs and sputum analysis using ZN analysis in the laboratory…”* Clinical Officer.

It is important to note that some health workers mentioned a cough lasting over 3 weeks, which is contrary to the guidelines from the National TB and Leprosy Program (NTLP). 276 NTLP recommends 2 instead of 3 weeks. The other symptoms mentioned were evening fevers, night sweats, weight loss and loss of appetite.

*“…. we normally diagnose TB after 3 weeks of a patient coughing, we tell the patients that they could have TB, then we find out if the patient has any of these symptoms; loss of appetite, evening fevers and sweating in the night. We tell the patient to go for a laboratory test, sputum analysis”* Enrolled Nurse.

*“…when they (patients) come to OPD, screening for tuberculosis is done. TB suspects are those who have had a prolonged cough longer than 3 weeks, unexplained fevers, night sweats and weight loss…”* Clinical Officer

## Discussion

Our study of the facilitators, barriers and health workers’ practices regarding the use of the ICF tool among health care workers in Jinja Municipality in Eastern Uganda shows that the

ICF tool is easy to use. However, barriers such as other competing tools, availability and lack of awareness and knowledge about the ICF tool affected its optimal use.

The ICF tool was found to be easy to use mainly due to its brevity and simplicity with clear instructions. The National and Leprosy Programs should capitalize on these advantages in its design to increase the uptake of the ICF tool.

However, because the ICF tool is presumed to be easy to use, comprehensive training has been overlooked and emphasis placed on support supervision. As a result, most of the health workers do not seem to understand the importance of using the ICF tool in TB screening.

The laboratory personnel were found to have slightly limited knowledge as regards to ICF. We recommend across-the-board training that should target everybody likely to encounter a TB patient. Despite the Laboratory staff usually only running samples, they also come in contact with patients. They could screen those referred for other tests but may have presumptive signs and symptoms for TB. Especially when they have been referred to the laboratory serially.

Health care workers raised various reasons that hindered the use of the ICF tool; these included other competing tools, the tools getting lost after screening since they are stand-alone and stock-outs of the tool were frequently mentioned. All the above barriers can be overcome by embedding the ICF tool into the patient clinical encounter forms.

This is the first study of its kind to assess the use of ICF tool in Uganda; the data presented will have national-level significance and application. The increased use of the ICF tool is expected to have a marked rise in the Case Notification Rate (CNR) of TB patients [5,12,13] with patients diagnosed earlier thereby reducing their mortality as well as eventually decreasing the transmission of disease in the community [5,13,14]. However, despite this studies several strengths, it had some limitations, this study had a relatively small sample size (N=24) and saturation may not have been reached.

## Conclusion

The ICF tool was considered to be easy to use by health workers and this facilitated their use. However, a variety of barriers continue to hamper its optimal use including other competing tools, availability, lack of awareness and knowledge about the ICF tool. This explains the low uptake of the ICF tool. To increase its utilization, there is a need to capitalize on the facilitators of the tool while getting ways of mitigating the barriers. There is also a need to improve awareness among health workers about the ICF tool, its importance and when and where it should be used.

## Data Availability

It is a qualitative study, there is no data available.

## Acknowledgement

This work is a product of a thesis by KM who completed his Masters of Public Health at Uganda Christian University, Mukono, Uganda. EAO was the graduate research supervisor for KM and is a doctorate research fellow at Makerere University College of Health Sciences, Kampala, Uganda as well as tutor at the University of London, UK. EAO received support to complete this work from the Uganda-Case Western Reserve University Research Collaboration via the Fogarty International Clinical Research Scholars and Fellows Program at Vanderbilt University and the American Relief and Recovery Act, Fogarty International Centre, National Institutes of Health, USA based at the JCRC Uganda (R24 TW007988).

## Author Contributions

The following authors participated in developing the idea into a concept (KM, EAO), data collection and analysis (KM, EAO), drafting the manuscript (KM), reviewing, critically 396 appraising and approving the final version of the manuscript (KM, EAO, JM).

